# Is life-course neighbourhood deprivation associated with frailty and frailty progression from age 70 to 82 in the Lothian Birth Cohort 1936?

**DOI:** 10.1101/2021.09.03.21263087

**Authors:** Gergő Baranyi, Miles Welstead, Janie Corley, Ian J. Deary, Graciela Muniz-Terrera, Paul Redmond, Niamh Shortt, Adele Taylor, Catharine Ward Thompson, Simon Cox, Jamie Pearce

**Affiliations:** Centre for Research on Environment, Society and Health, School of GeoSciences, The University of Edinburgh, Edinburgh, UK; Lothian Birth Cohorts, Department of Psychology, University of Edinburgh, Edinburgh, UK; Edinburgh Dementia Prevention, University of Edinburgh, Edinburgh, UK; OPENspace research centre, University of Edinburgh, Edinburgh, UK

**Keywords:** ageing, life-course approach, frailty, frailty progression, neighbourhood social deprivation, structured life-course modelling

## Abstract

**Background:** Neighbourhood features have been postulated as key predictors of frailty. However, evidence is mainly limited to cross-sectional studies without indication of long-term impact and developmental timing of the exposures. This study explored how neighbourhood social deprivation (NSD) across the life course is associated with frailty and frailty progression among older Scottish adults.

**Methods:** Participants (n=323) were from the Lothian Birth Cohort 1936 with historical measures of NSD in childhood (1936-1955), early adulthood (1956-1975) and mid-to-late adulthood (1976-2014). Frailty was measured five times between the ages of 70 and 82 years using the Frailty Index. Confounder-adjusted life-course models were assessed using a structured modelling approach with least angle regression; associations were estimated for frailty at baseline using linear regression, and for frailty progression using linear mixed-effects models.

**Results:** Accumulation was the most appropriate life-course model for males; greater accumulated NSD was associated with higher frailty at age 70 (b=0.017; 95%CI: 0.005, 0.029; *P*=0.007) with dominant exposure times in childhood and mid-to-late adulthood. Among females, mid-to-late adulthood sensitive period was the best-fit life-course model and higher NSD in this period was associated with widening frailty trajectories between age 70 and 82 (b=0.005; 95%CI: 0.0004, 0.009, *P*=0.033).

**Conclusions:** This is the first investigation of the life-course impact of neighbourhood deprivation on frailty in a cohort of older adults with residential information across their lives. Future research should explore neighbourhood mechanisms linking deprivation to frailty. Policies designed to address neighbourhood deprivation and inequalities across the full life course may support healthy ageing.

**Key messages:**

- Neighbourhood context might be associated with old-age frailty, but existing investigations are mainly based on cross-sectional data with limited understanding of the relative importance of exposure timing during the life course.
- Using a structured approach, we investigated how neighbourhood social deprivation across the life course is associated with frailty, and frailty progression, in a sample of older Scottish adults.
- Among males, accumulated neighbourhood social deprivation was moderately associated with frailty at age 70 but not with subsequent frailty trajectories; widening frailty trajectories between age 70 and 82 conditional on deprivation during mid-to-late adulthood were identified among females.
- Gendered experiences of living in deprived areas from childhood onwards may contribute to frailty which should be considered in policies supporting healthy ageing.

## INTRODUCTION

The world’s population is rapidly ageing, resulting in growing numbers of older adults and a greater proportion of the population aged 60+. Whereas population ageing is most advanced in high-income countries,^1^ changes to the demographic profile can be observed worldwide.^2^ In 2017, there were 962 million individuals aged 60+ which is expected to double by 2050 with an anticipated threefold rise among the oldest old (>80 years).^2^ Not everybody ages in the same way: age-related decline in physical and mental capacities and functional abilities are not homogenous.^3^

Frailty is an age-related syndrome characterised as increased vulnerability, loss of resistance to stressors, and decreased reserves of capacity, which develops as a consequence of cumulative declines in several inter-related physiological systems.^4,5^ Frailty among older people heightens the risk of falls,^6^ morbidity^4^ and mortality;^7^ it is linked to increased health- and social-care costs.^8^ The prevalence of frailty varies considerably, with higher rates among females,^9,10^ socioeconomically disadvantaged and ethnic minority groups.^8^ Similarly, a range of sociodemographic (e.g. socioeconomic status) and ‘lifestyle’ factors (e.g. physical activity) are associated with frailty progression.^11^

Despite growing interest in the contextual determinants of various aspects of health and well-being,^12,13^ there is an important research gap in understanding how environments – particularly local, neighbourhood-level factors – contribute to frailty.^3^ A recent review^3^ found that social and physical characteristics of neighbourhoods, including deprivation, ethnic diversity/heterogeneity, social cohesion or walkability are associated with frailty. However, the evidence relied heavily on cross-sectional data and none of the identified investigations utilised repeated neighbourhood assessments through time.^3^ Similarly, little is known about how neighbourhood-level factors are associated with frailty progression.^3,11^ Places evolve over time (e.g. urban redevelopment, economic decline); depending on the timing of exposure during human development, neighbourhood features may have a differential and long-lasting impact on health.^12^ Describing how living in different contexts across the life course is associated with frailty is crucial in identifying modifiable risk factors, understanding age-related decline and developing age-friendly policies to support healthy ageing.

Using rarely-available longitudinal individual- and area-level data, our study aims to fill this research gap by applying the life-course framework. First, using a structured approach,^14^ we identified the most appropriate life-course models for frailty at age 70. We considered sensitive periods (i.e. whether the association between neighbourhood and frailty is stronger in particular developmental periods), accumulation (i.e. whether the sum of exposures over time is associated with frailty), and effect modification (i.e. whether the impact of neighbourhood on frailty is modified by exposure at an earlier period). Second, we estimated the association between best-fit model(s) and frailty at age 70, and frailty progression between age 70 and 82.

## METHODS

### Study sample

Data were drawn from the Lothian Birth Cohort 1936 (LBC1936), which is a follow up study of the Scottish Mental Survey 1947 testing the intelligence of almost every child born in 1936 and attending schools in Scotland on June 4, 1947 (N=70,805).^15^ Between 2004 and 2007, surviving participants residing in Edinburgh and the Lothian region of Scotland were retraced and invited to participate in LBC1936 (mean age 70 years).^15^ The cohort (n=1091 at wave 1) has been followed up at age 73 (n=866), 76 (n=697), 79 (n=550) and 82 (n=431), with attrition mainly due to withdrawal (including from illness) and death.^16^

### Measures

#### Neighbourhood social deprivation

Historical residential addresses were collected retrospectively for the LBC1936 as part of a ‘life grid’ questionnaire administered in 2014 (mean age 78). With major historic events (e.g. the Falklands War of 1982) serving as memory prompts, surviving participants were asked to recall their home addresses for every decade of their lives.^12,16^ Out of 704 contacts, 593 provided 7423 addresses, which were geocoded using automatic geocoders (i.e. Nominatim, Google) and historical building databases.^12^

Neighbourhood social deprivation (NSD) for the City of Edinburgh was captured once a decade during participants’ life. Between 1931 and 1971 this was done with a historical index of multiple deprivation (i.e. population density, overcrowding, infant mortality, tenure, and amenities),^12^ and between 1981 and 2011 with the Carstairs index of deprivation (i.e. male unemployment, overcrowding, car ownership, and social class).^17^ All data were aggregated to 1961 census ward boundaries (n=23) to ensure consistent spatial units; comparability across decades was maintained by computing deprivation indices z-scores.^12^ We explored correlations between NSD scores (Supplementary Figure 1) and computed average exposure in childhood (1936-1955; age 0-19), young adulthood (1956-1975; age 20-39) and mid-to-late adulthood (1976-2014; age 40-78); these were computed for participants for whom at least one Edinburgh-based address was reported in all periods.

#### Frailty Index

Frailty was assessed across all follow-up waves utilising the Frailty Index, a continuous measure representing frailty as an accumulation of health deficits (e.g. symptoms, diagnosis, impairments) across multiple body systems.^18,19^ We extracted information for 30 variables covering physical, psychological and cognitive systems routinely collected for LBC1936 waves.^16^ Cut-off scores representing a deficit in each of the 30 frailty variables were previously established^20^ based on recommendations^18^ (see Supplementary Table 1). Frailty scores were calculated by summing each participant’s deficits and dividing by the total number of possible deficits (n=30). The indicator ranged between 0 and 1 with higher values showing higher degrees of frailty.

#### Covariates

Life-course covariates were identified based on the literature^12,20,21^ and presented in a directed acyclic graph (DAG) taking into consideration time-specific confounding, and selection into similar neighbourhoods (Figure 1). Variables included sex, age in years (time-variant), parental occupational social class (professional-managerial [I/II] versus skilled, partly skilled and unskilled [III/IV/V]),^22^ childhood IQ score at age 11 measured with the Moray House Test,^15^ years spent in full-time education, and childhood smoking (initiating before age 16). Adult OSC (I/II versus III/IV/V)^22^ and current smoking status (yes, no) were extracted at age 70.

**Figure 1.**
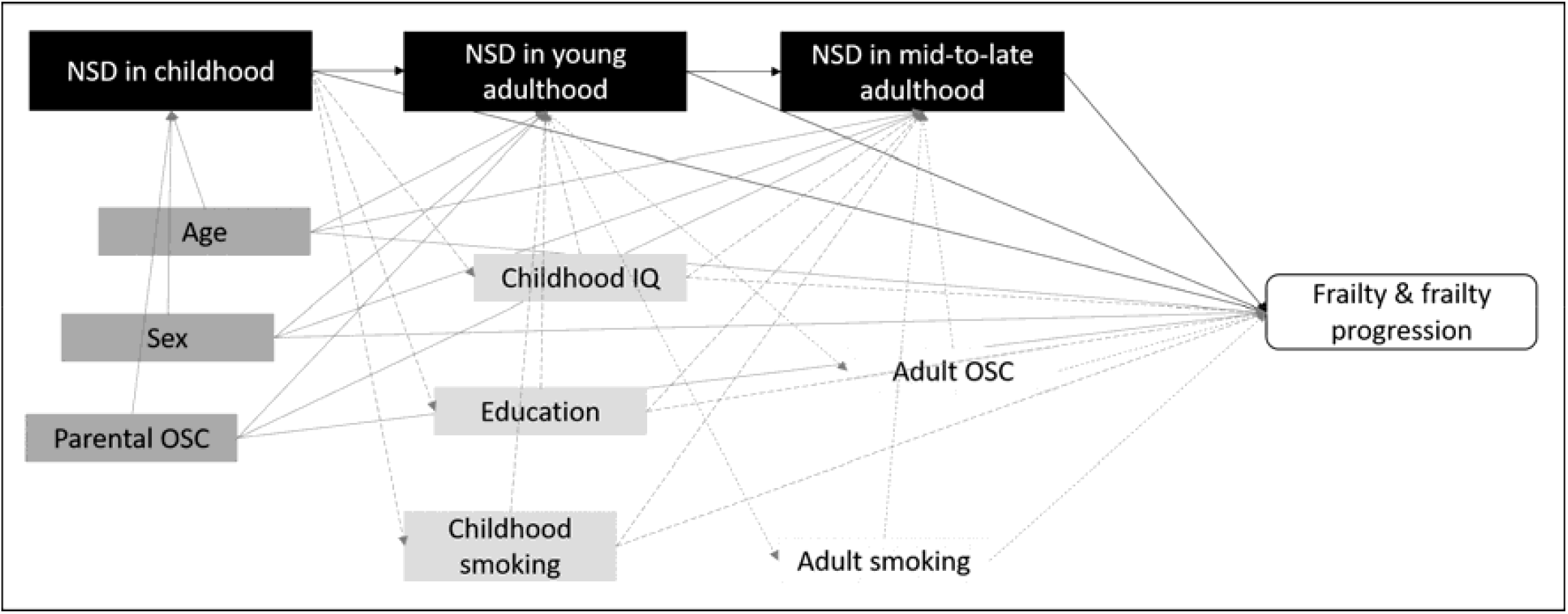
Directed acyclic graph presenting theoretical associations between neighbourhood social deprivation, frailty and selected life course covariates. Links between covariates are not shown for simplicity. Differently coloured covariates and arrows indicate time-specific confounding (i.e. dark grey – childhood onwards; medium grey – young adulthood onwards; light grey – mid-to-late adulthood). NSD - neighbourhood social deprivation; OSC – occupational social class.

### Statistical analysis

We compared study variables between included participants and the rest of the baseline sample to investigate sample bias, with differences tested using two-sample t-tests and chi-squared tests. To explore the life-course associations between NSD, frailty and frailty progression, we applied the modified two-stage structured life course modelling approach^14^ originally proposed by Mishra.^23^ Analyses were conducted in R 4.0.3,^24^ separately for men and women.^9-11^

In stage-one, the most appropriate life-course models for frailty at age 70 were identified using the Least Angle Regression (LARS) algorithm for variable selection. LARS provides a structured and unbiased way to select an input variable (or a combination) from multiple simultaneously competing ones with the strongest association to the outcome.^14^ It implements the least absolute shrinkage and selection operator (lasso) to identify the best-fit variable;^14,25^ after the first variable is selected, the procedure identifies the combination of two variables explaining the largest outcome variance, and so on until all input variables are selected. Following recommended practice,^26^ we used the elbow plot depicting the proportion of outcome variance (R^2^) explained by selected variable(s)^14^ and the covariance test for the lasso indicating improvement in explained outcome proportion (*P*<0.05).^27^

Six competing life-course models were encoded as LARS input variables. Sensitive periods were captured as NSD in childhood, young adulthood, and mid-to-late adulthood; accumulation was the average exposure across these. Effect modifications in early and later life were operationalised as interactions between childhood and young adulthood, and between young and mid-to-late adulthood NSD. To account for confounding, we regressed input variables on covariates identified as common confounders across all life-course models (age, parental OSC), and entered the model residuals into LARS.^14^

In stage-two, we estimated the effect size of selected models in a multiple regression framework separately for age 70 frailty and frailty progression between age 70 and 82. Three sets of confounders were considered relevant for the proposed life course models (see DAG):

- **Model 1**: age, parental OSC – common confounders for all life-course models and considered as most appropriate adjustment for childhood sensitive period.
- **Model 2**: Model 1 + years of education, childhood smoking, age 11 IQ – relevant confounders for young adulthood sensitive period and early life effect modification.
- **Model 3**: Model 2 + adult OSC, current smoking – relevant for accumulation, mid-to-late adulthood sensitive period and later life effect modification.

Where applicable, we also added NSD from the previous life-course period to account for selection into similar neighbourhoods.^28^ Calculations for frailty were based on linear regression. Frailty progression was fitted in linear mixed-effects regression with random intercepts and slopes – chosen as best fitting model – where NSD × age interaction represented change in frailty scores. Coefficients (b) with 95% confidence intervals (CI) based on scaled and mean-centred continuous variables were calculated; we also reported fully standardized coefficients (i.e. predictor and outcome) to aid interpretation of effect sizes (β). After fitting linear mixed-effects models for frailty progression, we calculated Johnson-Neyman intervals with adjustment for false discovery rates to explore where NSD slopes change between regions of significance and non-significance conditional on age. Given the limitation of our overall framework (i.e. sex-stratified analyses), we performed confirmatory analyses by testing the sex × NSD interaction in the total sample.

We ran four sets of sensitivity analyses. First, instead of using common confounders (i.e. age, parental OSC) to produce model residuals for LARS, we regressed life-course models on their specific confounders. Second, instead of including participants with at least one Edinburgh-based address in childhood, young adulthood and mid-to-late adulthood, we rerun models with individuals remaining in Edinburgh throughout their lives. Third, we considered NSD measures only until 2005, to avoid temporal overlap between exposure and outcome assessment (2004/2007-2017/2019). Last, we tested linearity of associations by replacing continuous NSD variables with categorical ones indicating low, moderate, and high deprivation.

## RESULTS

We included 323 individuals into our analyses; 30% of non-participants did not have Edinburgh-based addresses (Figure 2). Included participants were on average younger, and less likely to smoke or be frail at age 70. In the analytical sample, there were 161 men and 162 women; women had a higher IQ at age 11 and less likely to smoke before age 16. Participants’ exposure to socially deprived neighbourhoods decreased during the course of their lives, while their frailty increased across waves (Table 1).

**Figure 2.**
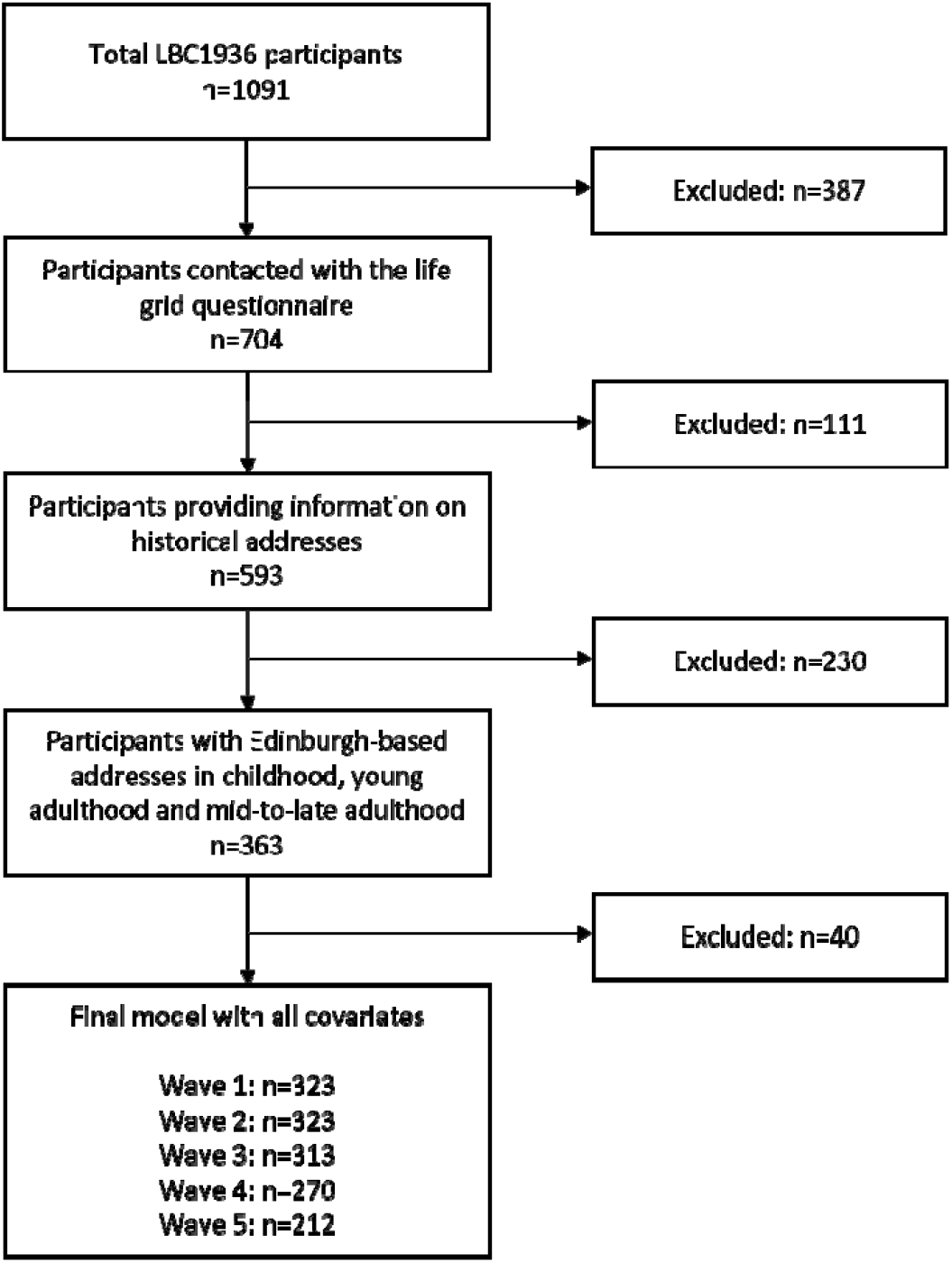
Flowchart indicating sample selection

**Table 1.** Descriptive statistics for included and excluded participants, Lothian Birth Cohort 1936

|  | **Total**<br>**LBC1936**<br>**(n=1091)** | **Excluded**<br>**(n=768)** | **Included**<br>**(n=323)** | ***P*-**<br>**value** | **Male**<br>**(n=161)** | **Female**<br>**(n=162)** | ***P*-value** |
| --- | --- | --- | --- | --- | --- | --- | --- |
| Age at baseline, mean (SD) | 69.53 (0.83) | 69.62 (0.86) | 69.34 (0.74) | <0.001 | 69.33 (0.75) | 69.35 (0.73) | 0.853 |
| Sex, n (%) |  |  |  |  |  |  |  |
| Male | 548 (50.23%) | 387 (50.39%) | 161 (49.85%) |  |  |  |  |
| Female | 543 (49.77%) | 381 (49.61%) | 162 (50.15%) | 0.922 |  |  |  |
| Parental occupational social class, n (%) |  |  |  |  |  |  |  |
| I and II | 260 (27.08%) | 172 (22.40%) | 88 (27.24%) |  | 44 (27.33%) | 44 (27.16%) |  |
| III, IV and V | 700 (72.92%) | 465 (60.55%) | 235 (72.76%) | 0.997 | 117 (72.67%) | 118 (72.84%) | 1.000 |
| IQ at age 11, mean (SD) | 100.00 (15.00) | 99.44 (15.31) | 101.23 (14.22) | 0.068 | 99.56 (15.01) | 102.89 (13.23) | 0.035 |
| Years spent in education, mean (SD) | 10.74 (1.13) | 10.78 (1.16) | 10.64 (1.06) | 0.058 | 10.66 (1.04) | 10.62 (1.07) | 0.727 |
| Childhood smoking (≤16 years), n (%) |  |  |  |  |  |  |  |
| Yes | 226 (20.71%) | 166 (21.61%) | 60 (18.58%) |  | 44 (27.33%) | 16 (9.88%) |  |
| No | 865 (79.29%) | 602 (78.39%) | 263 (81.42%) | 0.294 | 117 (72.67%) | 146 (90.12%) | <0.001 |
| Adult occupational social class, n (%) |  |  |  |  |  |  |  |
| I and II | 592 (55.33%) | 413 (53.78%) | 179 (55.42%) |  | 82 (50.93%) | 97 (59.88%) |  |
| III, IV and V | 478 (44.67%) | 334 (43.49%) | 144 (44.58%) | 1.000 | 79 (49.07%) | 65 (40.12%) | 0.132 |
| Current smoking at baseline, n (%) |  |  |  |  |  |  |  |
| Yes | 125 (11.46%) | 101 (13.15%) | 24 (7.43%) |  | 10 (6.21%) | 14 (8.64%) |  |
| No | 966 (88.54%) | 667 (86.85%) | 299 (92.57%) | 0.009 | 151 (93.79%) | 148 (91.36%) | 0.535 |
| Frailty Index, mean (SD) |  |  |  |  |  |  |  |
| wave 1 | 0.16 (0.09) | 0.17 (0.09) | 0.14 (0.07) | <0.001 | 0.14 (0.08) | 0.15 (0.07) | 0.757 |
| wave 2 |  |  |  |  | 0.17 (0.08) | 0.17 (0.07) | 0.568 |
| wave 3 |  |  |  |  | 0.20 (0.08) | 0.20 (0.08) | 0.950 |
| wave 4 |  |  |  |  | 0.21 (0.09) | 0.21 (0.09) | 0.908 |
| wave 5 |  |  |  |  | 0.21 (0.09) | 0.21 (0.09) | 0.833 |
| Neighbourhood social deprivation, mean (SD) |  |  |  |  |  |  |  |
| Childhood |  |  |  |  | 0.59 (3.45) | 0.24 (3.13) | 0.337 |
| Young adulthood |  |  |  |  | -0.49 (2.78) | -0.79 (2.42) | 0.303 |
| Mid-to-late adulthood |  |  |  |  | -1.99 (2.87) | -1.91 (2.70) | 0.797 |

*P*-values are based on two-sample *t*-tests for mean difference, and on chi-squared tests for differences in distribution. Abbreviations: SD – standard deviation.

Among males, the LARS procedure identified accumulation as the best-fit life-course model, accounting for 7.16% of the unexplained variance (*P*<0.001). Although the elbow plot (Figure 3) indicated further improvements by adding mid-to-late adulthood, and childhood sensitive periods (R^2^=0.134), these steps were not supported by the covariance test of lasso (*P*>0.05). Among females, mid-to-late adulthood sensitive period was the first selected model (R^2^=0.022); however, it was not supported by the covariance test (*P*=0.087).

**Figure 3.**
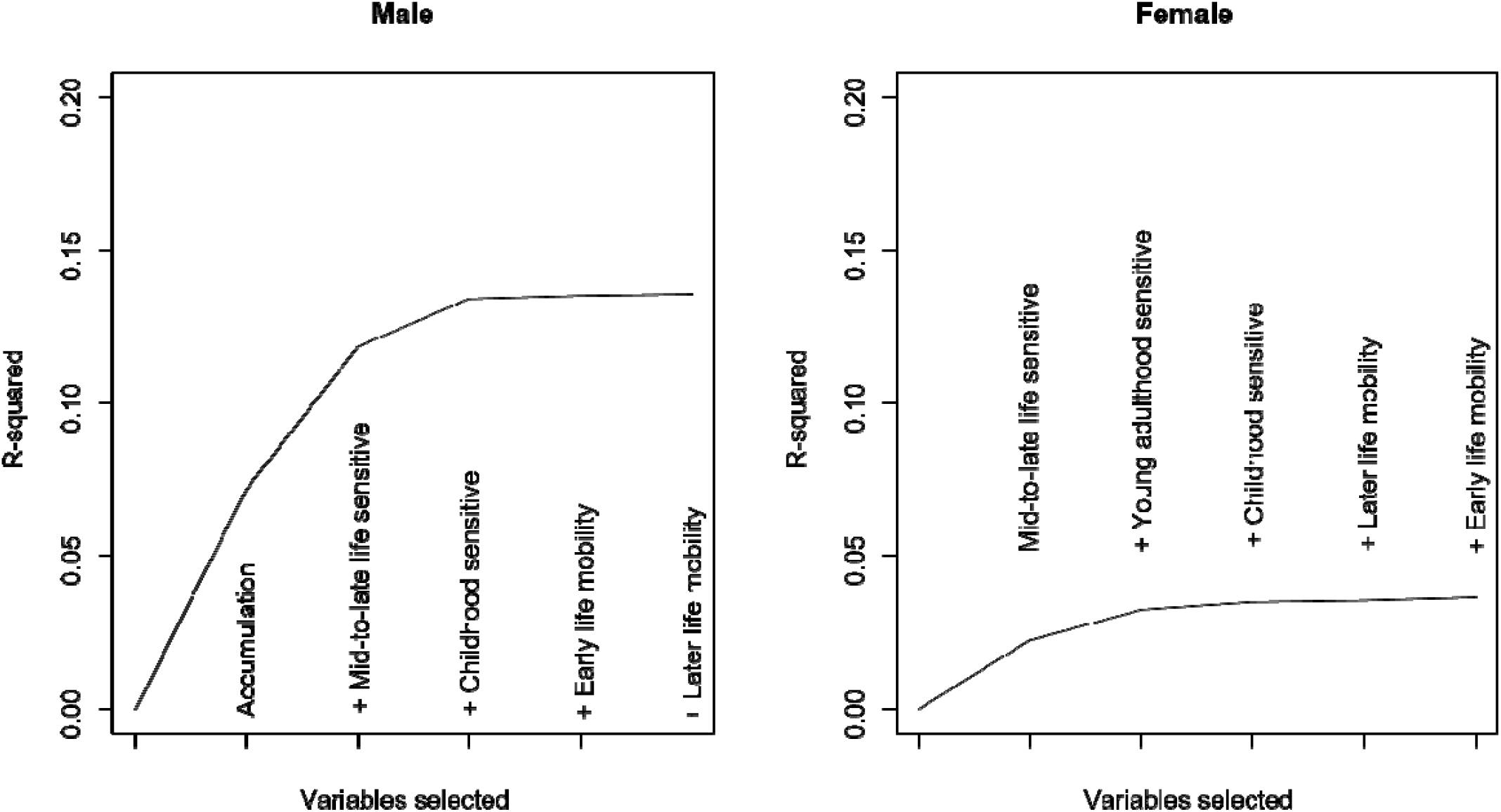
Elbow plot illustrating LARS selection procedure for life-course models of neighbourhood social deprivation based on explained outcome variance (i.e. frailty at age 70). LARS input variables were residuals of variable-encoded life course models, regressed on age and parental occupational social class as common confounders of all life-course models. The LARS procedure first selects the variable with the largest outcome variance explained, followed by a combination of additional variables with increasingly strong associations to the outcome. Young adulthood sensitive period for male, accumulation for female were dropped because of collinearity.

After choosing the best-fit life course models, we first estimated the association between NSD and frailty at baseline. For both selected models, full adjustment models (Model 3) were deemed most appropriate based on our DAG. Among men, a 1 SD higher score in accumulated NSD was associated with a 0.017 (95%CI: 0.005, 0.029; *P*=0.007) higher value in the Frailty Index score at age 70, presenting a moderate effect size (β=0.223) (Table 2). *Post-hoc* linear regression adjusted for false discovery rate explored periods most likely contributing to accumulation: childhood (Model 1: b=0.021; 95%CI: 0.009, 0.031, *P-adjusted=*0.027) and mid-to-late adulthood NSD (Model 3: b=0.015, 95%CI: 0.002, 0.027, *P-adjusted*=0.029) were associated with frailty, pointing towards relaxed accumulation (i.e. periods are not contributing equally to the risk) among men.^29^ Among females, mid-to-late adulthood NSD was not associated with frailty at baseline (b=0.010; 95%CI:-0.002, 0.022; *P*=0.109). In the total sample, sex-differences were present for all reported associations (*P*<0.05).

**Table 2.** Associations between neighbourhood social deprivation, frailty and frailty progression in the Lothian Birth Cohort 1936

|  | Male (n=161) |  |  |  | Female (n=162) |  |  |  |
| --- | --- | --- | --- | --- | --- | --- | --- | --- |
|  | Accumulation |  |  |  | Mid-to-late adulthood sensitive period<sup>a</sup> |  |  |  |
|  | b | 95% CI | P-value | β | b | 95% CI | P-value | β |
| **Frailty at age 70 (wave 1)** |  |  |  |  |  |  |  |  |
| Neighbourhood social deprivation | 0.017 | 0.005, 0.029 | 0.007 | 0.223 | 0.010 | -0.002, 0.022 | 0.109 | 0.144 |
| Age in years | 0.008 | -0.002, 0.019 | 0.121 | 0.113 | 0.012 | 0.001, 0.022 | 0.027 | 0.174 |
| Parental OSC (ref: I & II) | -0.018 | -0.044, 0.007 | 0.155 | -0.246 | -0.001 | -0.024, 0.023 | 0.963 | -0.008 |
| IQ at age 11 | -0.009 | -0.021, 0.002 | 0.108 | -0.126 | -0.018 | -0.029, 0.007 | 0.002 | -0.263 |
| Years spent in education | -0.020 | -0.034, -0.006 | 0.005 | -0.269 | 0.001 | -0.011, 0.013 | 0.907 | 0.010 |
| Childhood smoking (ref: no) | 0.011 | -0.013, 0.035 | 0.363 | 0.146 | 0.002 | -0.032, 0.037 | 0.893 | 0.035 |
| Adult OSC (ref: I & II) | 0.007 | -0.017, 0.030 | 0.583 | 0.088 | -0.018 | -0.040, 0.005 | 0.117 | -0.265 |
| Current smoking (ref: no) | -0.009 | -0.052, 0.034 | 0.675 | -0.122 | 0.004 | -0.032, 0.040 | 0.818 | 0.062 |
| **Frailty progression between age 70 and 82 (wave 1-5)** |  |  |  |  |  |  |  |  |
| *Fixed effects* |  |  |  |  |  |  |  |  |
| Neighbourhood social deprivation | 0.017 | 0.005, 0.029 | 0.005 | 0.195 | 0.016 | 0.005, 0.027 | 0.004 | 0.196 |
| Age in years | 0.029 | 0.024, 0.034 | 0.000 | 0.333 | 0.027 | 0.023, 0.032 | 0.000 | 0.328 |
| Parental OSC (ref: I & II) | -0.014 | -0.038, 0.010 | 0.260 | -0.158 | 0.007 | -0.014, 0.029 | 0.509 | 0.087 |
| IQ at age 11 | -0.009 | -0.020, 0.001 | 0.092 | -0.106 | -0.022 | -0.032, 0.012 | 0.000 | -0.264 |
| Years spent in education | -0.018 | -0.031, -0.005 | 0.008 | -0.208 | 0.001 | -0.010, 0.011 | 0.922 | 0.006 |
| Childhood smoking (ref: no) | 0.014 | -0.008, 0.036 | 0.213 | 0.162 | 0.005 | -0.025, 0.036 | 0.728 | 0.066 |
| Adult OSC (ref: I & II) | 0.004 | -0.019, 0.026 | 0.743 | 0.043 | -0.023 | -0.043, 0.003 | 0.023 | -0.282 |
| Current smoking (ref: no) | -0.003 | -0.043, 0.038 | 0.899 | -0.030 | 0.008 | -0.025, 0.040 | 0.639 | 0.093 |
| Neighbourhood social deprivation*Age | -0.001 | -0.006, 0.004 | 0.645 | -0.013 | 0.005 | 0.0004, 0.009 | 0.033 | 0.058 |
| *Random effects* |  |  |  |  |  |  |  |  |
| Intercept, variance (SD) | 0.136 | (0.368) |  |  | 0.105 | (0.323) |  |  |
| Slope, variance (SD) | 0.000 | (0.005) |  |  | 0.000 | (0.005) |  |  |
| Residuals, variance (SD) | 0.001 | (0.037) |  |  | 0.001 | (0.038) |  |  |

*Note:* Regression coefficients (b) and 95% confidence intervals (CI) are displayed; we also provide fully standardized coefficients (β) to aid interpretation. Multivariate models are based on linear regression for frailty at age 70 and on linear mixed-effects regression with random intercepts and slopes for frailty progression; continuous predictors are mean centred and scaled.
Abbreviations: OSC – occupational social class, SD – standard deviation
<sup>a</sup> Models are adjusted for neighbourhood social deprivation in the previous developmental period (i.e. young adulthood).

Using linear mixed-effects regression, we investigated NSD × age interaction on frailty progression. We found that frailty trajectories were not associated with accumulated NSD in males (b=-0.001; 95%CI: -0.006, 0.004, *P*=0.645). However, in females, a 1 SD higher score in mid-to-late adulthood NSD was associated with 0.005 (95%CI: 0.0004, 0.009, *P*=0.033) change in Frailty Index score for each 1 SD increase in age (Table 2; Figure 4); indicating widening NSD-based inequalities in frailty levels between age 70 and 82 (β=0.058). Sex differences were confirmed in the total sample (*P*<0.05). Finally, Johnson-Neyman intervals indicated that the association with NSD first materialises at age 70.5 among females; but they also suggested that the association among males might diminish after the age of 81.2 (Supplementary Figure 2).

**Figure 4.**
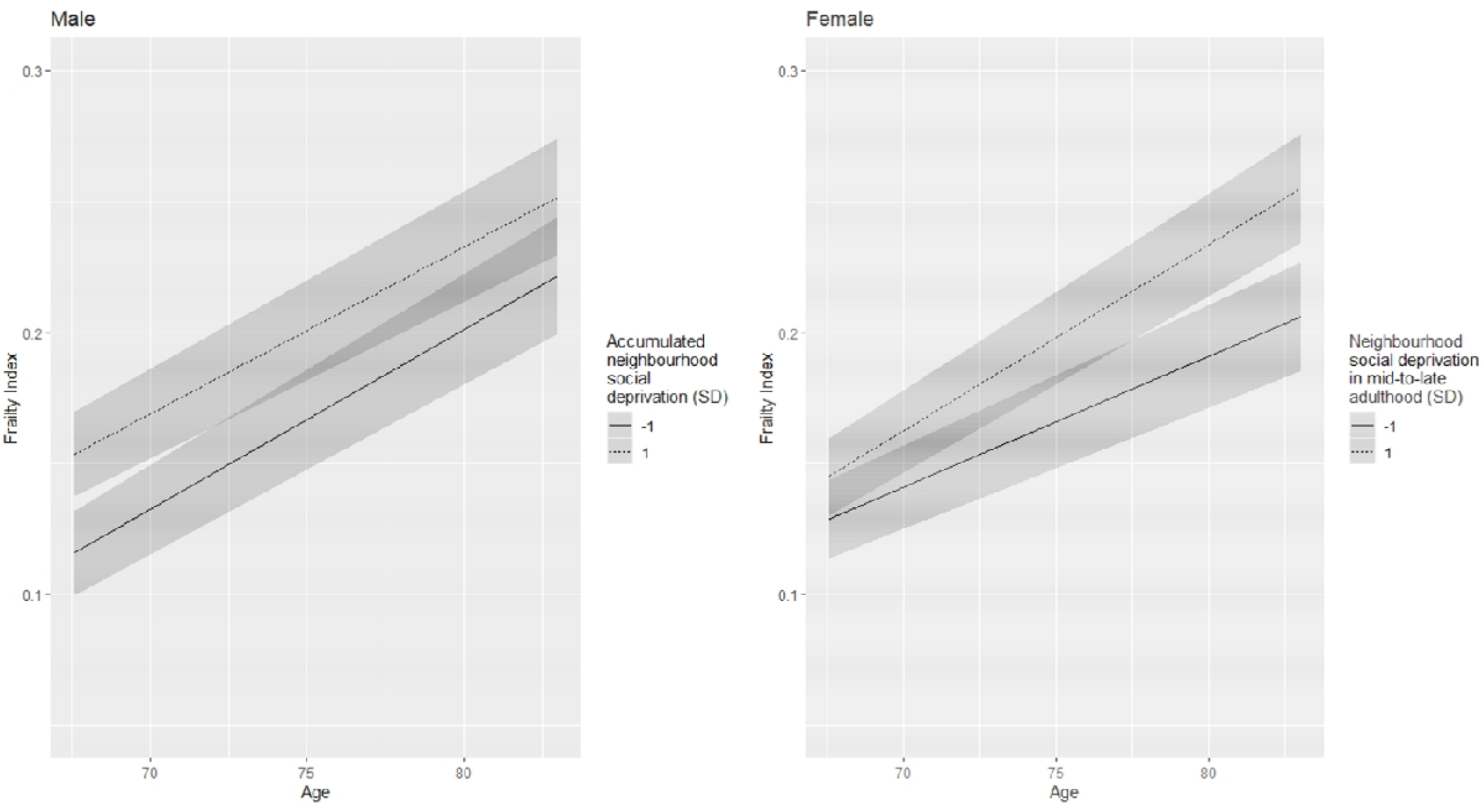
Frailty progression among males and females in the Lothian Birth Cohort 1936 by selected neighbourhood social deprivation life-course models. Plots present predicted probabilities with discrete predictors held constant at their proportions. Calculations are based on most appropriate life-course models for male (i.e. accumulation) and female (i.e. mid-to-late adulthood sensitive period) participants. Abbreviation: SD – standard deviation

Sensitivity analyses extended and confirmed our findings. When we reran LARS model selection with regression residuals adjusted for all relevant DAG-based life course-specific confounders, we found that childhood (*P*=0.006) and mid-to-late adulthood (*P*=0.022) sensitive periods were most appropriate models for male (R^2^=0.095). Among female, mid-to-late adulthood sensitive period remained as first selected (R^2^=0.003; *P*=0.757). Stage-two results were robust for frailty at baseline but became non-significant for frailty progression when the sample was restricted to LBC1936 participants living in Edinburgh throughout their lives (Supplementary Table 2; n=247), or when temporal overlap between NSD and outcome assessment was eliminated (Supplementary Table 3; n=322). The latter results stressed the importance of contemporaneous NSD exposure in frailty progression among females. When replacing continuous exposures with tertiles, findings confirmed linear relationships (Supplementary Table 4).

## DISCUSSION

Neighbourhood social deprivation is an important predictor of frailty and frailty progression in old age, but the life-course relationship differs by sex. Using a structured modelling approach, we identified the relaxed accumulation hypothesis as best capturing the link between NSD and frailty among males, whereby the impact of living in socially deprived areas in childhood and in mid-to-late adulthood contributed to higher frailty in older age. Among females, higher NSD in mid-to-late adulthood was associated with faster frailty progression: divergent slopes first materialised around age 70.5.

Consistent with evidence relating to different health outcomes,^21,28,30^ neighbourhood deprivation in mid-to-late adulthood was associated with frailty and its progression, in addition to the impact of early exposures among males. Structural differences across neighbourhoods, including social deprivation, can be linked to frailty via the stress pathway or through variations in providing collective resources and opportunities to residents to support their health and wellbeing.^31-34^ Living in socially deprived neighbourhoods may affect health and frailty, by accumulation of stress over time. A recent investigation demonstrated that long-term exposure to deprived neighbourhoods is associated with worse allostatic load: wear and tear on the body likely linked to chronic psychological stress exposure.^34^

Advantaged neighbourhoods may provide more opportunities to support health and wellbeing. Neighbourhood-based social processes (e.g. social cohesion, social participation) are protective against frailty by creating and maintaining social connections and support networks, and by buffering stress.^31,32,34,35^ Availability of recreational and cultural facilities has been associated with slower age-related decline.^36^ While perceiving residential areas as unsafe^32^ or deteriorating^31^ are associated with frailty, likely via higher stress levels, avoidance behaviour and maladaptive coping mechanisms,^31,37^ greater access to green space could stimulate engagement in physical and social activities, improving frailty status.^38^ However, understanding why neighbourhood-based inequalities in frailty persisted among males during almost the entire follow-up period (age 70-82) of the study but only first materialised at age 70.5 for females, needs further exploration, and might be linked to the role of neighbourhood resources and stressors across the life course.

In addition to mid-to-late adulthood exposure, deprivation in childhood was linked to frailty among males. Childhood is a formative developmental period; living in deprived neighbourhoods at an early age can adversely affect health in childhood and early adulthood,^39^ potentially through disrupting stress regulation^40^ or through alterations to the epigenome.^41^ Moreover, early life exposure can predict subsequent adverse (neighbourhood) exposures as described in the chains of risks hypothesis.^21^ Sex differences in early-life context may be linked to higher susceptibility and exposure to environmental influences among boys partly related to decreased parental supervision and stronger neighbourhood influences on future employment aspirations.^42^ Gendered early-life neighbourhood experiences were likely even more distinct in the first half of the 20^th^ century, with greater expectation of girls undertaking household domestic work while boys were more engaged in activities in their wider neighbourhood.

To our knowledge, this is the first study exploring the impact of neighbourhood context across most of the life course on frailty, and frailty trajectories. We utilised information on NSD covering the period from birth to late adulthood, repeated measures of frailty based on 30 health deficits, and key life-course confounders (e.g. childhood intelligence). Applying the novel structured life-course modelling approach reduced the risk of bias arising from simultaneously testing competing theoretical models and enabled us to choose parsimonious life course models, without overinflating effect size estimates during variable selection^25^ or biasing hypothesis test.^27^

Still, our study has several limitations. First, we had a modest sample size partly due to missing NSD measures for LBC1936 participants not residing in Edinburgh in key developmental periods. Second, residential addresses were collected retrospectively which is prone to recall bias.^12^ Third, information on NSD was aggregated at the ward-level and is limited to a small number of indicators that were not consistently available in official records throughout the study. Consequently, NSD was measured with two strongly correlated but distinct constructs.^12^ Fourth, although the analytical sample was similar to the full LBC1936 cohort in terms of socioeconomic indicators, it included healthier and younger participants further limiting the generalisability of findings. Last, as structural life-course modelling is not available for outcomes with repeated measurements, we ran LARS variable selection only for frailty at age 70. Future methodological developments would usefully take into account change in outcome levels.

## Conclusions

Our findings showed in an Edinburgh-based sample that neighbourhood social deprivation across the life course matters for frailty in older adulthood. While the impact of deprivation likely accumulates among males, with childhood and mid-to-late adulthood being pertinent, among females living in deprived areas during the second part of the life might be more relevant. Given above limitations, future research could usefully replicate our findings in large-scale longitudinal studies with more diverse populations, and explore specific neighbourhood mechanisms (e.g. social, environmental, geographic, institutional)^43^ linking structural area differences to age-related decline. Understanding causal routes by which individuals growing up, living and ageing in different context across their life course become frail and identifying vulnerable groups may have policy implication. Having access to good quality neighbourhoods from childhood onwards and placing multimodal frailty interventions^36^ in deprived areas may support healthy ageing by preventing and slowing age-related decline. Integrating the life-course framework into community-based policies likely present an opportunity to maintain health and wellbeing in the context of global population ageing.

### Ethics approval

The LBC1936 study was conducted according to the Declaration of Helsinki guidelines with ethical permission obtained from the Multi-Centre Research Ethics Committee for Scotland (MREC/01/0/56), Lothian Research Ethics Committee (wave 1, LREC/2003/2/29), and the Scotland A Research Ethics Committee (waves 2-5, 07/MRE00/58). Written consent was obtained from all participants.

## Supporting information

STROBE

## Data Availability

Data was obtained from the Lothian Birth Cohort 1936, more information can be found at https://www.lothianbirthcohort.ed.ac.uk/

## Funding

This work was supported by the Economic and Social Research Council, UK (grant award ES/P008585/1). The LBC1936 study is supported by Age UK (Disconnected Mind programme grant).

## Acknowledgements

The Lothian Birth Cohort 1936 study acknowledges the financial support of NHS Research Scotland (NRS), through Edinburgh Clinical Research Facility. We gratefully acknowledge the contributions of the LBC1936 participants and members of the LBC1936 research team who collect and manage the LBC data. We thank for Dr Andrew Smith for his guidance on the structured life-course modelling approach.

## Conflict of Interest

None declared.

## Notes

### Competing Interest Statement

The authors have declared no competing interest.

## REFERENCES

1. Rechel B, Grundy E, Robine JM, et al. Ageing in the European Union. Lancet 2013; 381(9874): 1312–22.

2. United Nations, Department of Economic and Social Affairs, Division. P. World Population Ageing 2017 - Highlights. New York: United Nations, 2017.

3. Fritz H, Cutchin MP, Gharib J, Haryadi N, Patel M, Patel N. Neighborhood Characteristics and Frailty: A Scoping Review. Gerontologist 2020; 60(4): e270–e85.

4. Clegg A, Young J, Iliffe S, Rikkert MO, Rockwood K. Frailty in elderly people. Lancet 2013; 381(9868): 752–62.

5. Fried LP, Tangen CM, Walston J, et al. Frailty in older adults: evidence for a phenotype. J Gerontol A Biol Sci Med Sci 2001; 56(3): M146–56.

6. Cheng MH, Chang SF. Frailty as a Risk Factor for Falls Among Community Dwelling People: Evidence From a Meta-Analysis. J Nurs Scholarsh 2017; 49(5): 529–36.

7. Kojima G, Iliffe S, Walters K. Frailty index as a predictor of mortality: a systematic review and meta-analysis. Age Ageing 2018; 47(2): 193–200.

8. Hoogendijk EO, Afilalo J, Ensrud KE, Kowal P, Onder G, Fried LP. Frailty: implications for clinical practice and public health. Lancet 2019; 394(10206): 1365–75.

9. Gordon EH, Peel NM, Samanta M, Theou O, Howlett SE, Hubbard RE. Sex differences in frailty: A systematic review and meta-analysis. Exp Gerontol 2017; 89: 30–40.

10. O’Caoimh R, Sezgin D, O’Donovan MR, et al. Prevalence of frailty in 62 countries across the world: a systematic review and meta-analysis of population-level studies. Age Ageing 2021; 50(1): 96–104.

11. Welstead M, Jenkins ND, Russ T, Luciano M, Muniz-Terrera G. A Systematic Review of Frailty Trajectories: Their Shape And Influencing Factors. Gerontologist 2020.

12. Pearce J, Cherrie M, Shortt N, Deary I, Ward Thompson C. Life course of place: A longitudinal study of mental health and place. Transactions of the Institute of British Geographers 2018; 43(4): 555–72.

13. Diez Roux AV, Mair C. Neighborhoods and health. Ann N Y Acad Sci 2010; 1186: 125–45.

14. Smith AD, Hardy R, Heron J, et al. A structured approach to hypotheses involving continuous exposures over the life course. Int J Epidemiol 2016; 45(4): 1271–9.

15. Deary IJ, Gow AJ, Pattie A, Starr JM. Cohort profile: the Lothian Birth Cohorts of 1921 and 1936. Int J Epidemiol 2012; 41(6): 1576–84.

16. Taylor AM, Pattie A, Deary IJ. Cohort Profile Update: The Lothian Birth Cohorts of 1921 and 1936. Int J Epidemiol 2018; 47(4): 1042–r.

17. UK Data Service. How the 2001 scores were calculated, n.d.

18. Searle SD, Mitnitski A, Gahbauer EA, Gill TM, Rockwood K. A standard procedure for creating a frailty index. BMC Geriatr 2008; 8(1): 24.

19. Mitnitski AB, Mogilner AJ, Rockwood K. Accumulation of deficits as a proxy measure of aging. ScientificWorldJournal 2001; 1: 323–36.

20. Welstead M, Muniz-Terrera G, Russ TC, et al. Inflammation as a risk factor for the development of frailty in the Lothian Birth Cohort 1936. Exp Gerontol 2020; 139: 111055.

21. Jivraj S, Murray ET, Norman P, Nicholas O. The impact of life course exposures to neighbourhood deprivation on health and well-being: a review of the long-term neighbourhood effects literature. Eur J Public Health 2020; 30(5): 922–8.

22. Office of Population Censuses and Surveys. Classification of Occupations and Coding Index. London, UK: HMSO, 1980.

23. Mishra G, Nitsch D, Black S, De Stavola B, Kuh D, Hardy R. A structured approach to modelling the effects of binary exposure variables over the life course. Int J Epidemiol 2009; 38(2): 528–37.

24. R Core Team. R: A language and environment for statistical computing. Vienna, Austria: R Foundation for Statistical Computing; 2021.

25. Efron B, Hastie T, Johnstone I, Tibshirani R. Least angle regression. The Annals of Statistics 2004; 32(2): 407-99, 93.

26. Marini S, Davis KA, Soare TW, et al. Adversity exposure during sensitive periods predicts accelerated epigenetic aging in children. Psychoneuroendocrinology 2020; 113: 104484.

27. Lockhart R, Taylor J, Tibshirani RJ, Tibshirani R. A Significance Test for the Lasso. Ann Stat 2014; 42(2): 413–68.

28. Jivraj S, Norman P, Nicholas O, Murray ET. Are there sensitive neighbourhood effect periods during the life course on midlife health and wellbeing? Health Place 2019; 57: 147–56.

29. Murray ET, Mishra GD, Kuh D, Guralnik J, Black S, Hardy R. Life course models of socioeconomic position and cardiovascular risk factors: 1946 birth cohort. Ann Epidemiol 2011; 21(8): 589–97.

30. Gustafsson PE, Bozorgmehr K, Hammarstrom A, Sebastian MS. What role does adolescent neighborhood play for adult health? A cross-classified multilevel analysis of life course models in Northern Sweden. Health Place 2017; 46: 137–44.

31. Caldwell JT, Lee H, Cagney KA. Disablement in Context: Neighborhood Characteristics and Their Association With Frailty Onset Among Older Adults. J Gerontol B Psychol Sci Soc Sci 2019; 74(7): e40–e9.

32. Cramm JM, Van Dijk HM, Nieboer AP. The creation of age-friendly environments is especially important to frail older people. Ageing and Society 2016; 38(4): 700–20.

33. Aranda MP, Ray LA, Snih SA, Ottenbacher KJ, Markides KS. The protective effect of neighborhood composition on increasing frailty among older Mexican Americans: a barrio advantage? J Aging Health 2011; 23(7): 1189–217.

34. Prior L. Allostatic Load and Exposure Histories of Disadvantage. Int J Environ Res Public Health 2021; 18(14): 7222.

35. Duppen D, Van der Elst MCJ, Dury S, Lambotte D, De Donder L D S. The Social Environment’s Relationship With Frailty: Evidence From Existing Studies. J Appl Gerontol 2019; 38(1): 3–26.

36. Rogers NT, Fancourt D. Cultural Engagement Is a Risk-Reducing Factor for Frailty Incidence and Progression. J Gerontol B Psychol Sci Soc Sci 2020; 75(3): 571–6.

37. Baranyi G, Di Marco MH, Russ TC, Dibben C, Pearce J. The impact of neighbourhood crime on mental health: A systematic review and meta-analysis. Soc Sci Med 2021; 282: 114106.

38. Yu R, Wang D, Leung J, Lau K, Kwok T, Woo J. Is Neighborhood Green Space Associated With Less Frailty? Evidence From the Mr. and Ms. Os (Hong Kong) Study. J Am Med Dir Assoc 2018; 19(6): 528–34.

39. Kivimaki M, Vahtera J, Tabak AG, et al. Neighbourhood socioeconomic disadvantage, risk factors, and diabetes from childhood to middle age in the Young Finns Study: a cohort study. Lancet Public Health 2018; 3(8): e365–e73.

40. Finegood ED, Rarick JRD, Blair C, Family Life Project I. Exploring longitudinal associations between neighborhood disadvantage and cortisol levels in early childhood. Dev Psychopathol 2017; 29(5): 1649–62.

41. Reuben A, Sugden K, Arseneault L, et al. Association of Neighborhood Disadvantage in Childhood With DNA Methylation in Young Adulthood. JAMA Netw Open 2020; 3(6): e206095.

42. Leventhal T, Brooks-Gunn J. The neighborhoods they live in: the effects of neighborhood residence on child and adolescent outcomes. Psychol Bull 2000; 126(2): 309–37.

43. Galster GC. The Mechanism(s) of Neighbourhood Effects: Theory, Evidence, and Policy Implications. In: van Ham M, Manley D, Bailey N, Simpson L, Maclennan D, eds. Neighbourhood Effects Research: New Perspectives. Dordrecht: Springer Netherlands; 2012: 23–56.

