## Supplement for "Is life-course neighbourhood deprivation associated with frailty and frailty progression from age 70 to 82 in the Lothian Birth Cohort 1936?"

**Supplementary Figure 1:** Heatmap depicting Pearson’s correlation coefficients between neighbourhood social deprivation scores across participants’ life course

**Supplementary Table 1:** Health deficits included in the Frailty Index

**Supplementary Figure 2:** Johnson-Neyman plots of neighbourhood social deprivation slopes and their confidence bands depicting regions of significance conditional on age

**Supplementary Table 2**: Main models among Lothian Birth Cohort 1936 participants living in Edinburgh during each decade of their lives

**Supplementary Table 3**: Main models based on non-overlapping exposure and outcome measurements in the Lothian Birth Cohort 1936

**Supplementary Table 4**: Main models with categorical neighbourhood social deprivation variables in the Lothian Birth Cohort 1936

**Supplementary Figure 1:** Heatmap depicting Pearson’s correlation coefficients between neighbourhood social deprivation scores across participants’ life course.

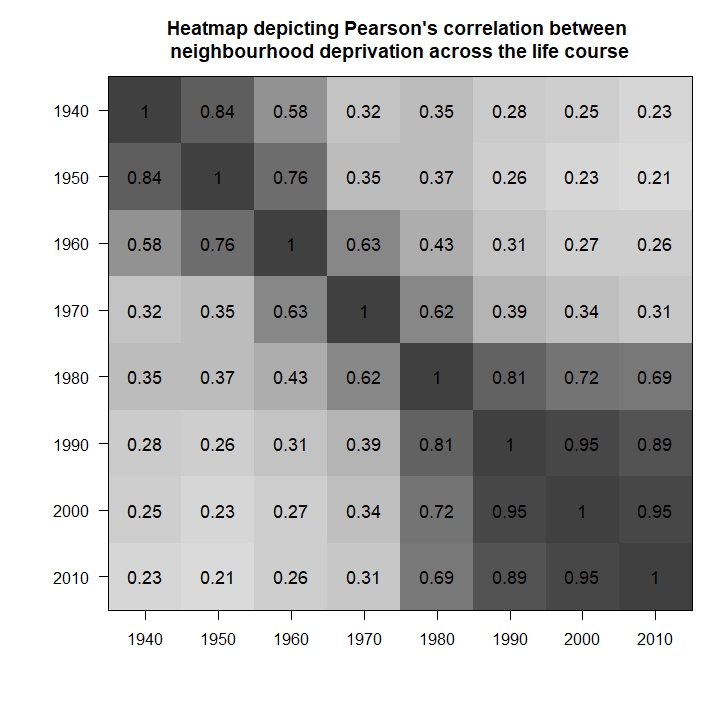

*Note*: correlation coefficients could be calculated only for participants providing Edinburgh-based addresses at least once for each decades of their lives (n=247).

**Supplementary Table 1:** Health deficits included in the Frailty Index.^a^

| **Items** | **Coding** | **Notes** |
| --- | --- | --- |
| Systolic blood pressure | <5th percentile (1), 5th-20th percentile (0.5), >20th percentile (0) | Recommended technique (Theou et al., 2015) |
| Diabetes (self-reported) | Yes (1) or No (0) | Already binary variable |
| High Cholesterol (self-reported) | Yes (1) or No (0) | Already binary variable |
| Heart problems (self-reported) | Yes (1) or No (0) | Already binary variable |
| Stroke or mini stroke (self-reported) | Yes (1) or No (0) | Already binary variable |
| Crampy pains in calves (self-reported) | Yes (1) or No (0) | Already binary variable |
| Blood circulation issues (self-reported) | Yes (1) or No (0) | Already binary variable |
| Thyroid Disorder (self-reported) | Yes (1) or No (0) | Already binary variable |
| Cancer (self-reported) | Yes (1) or No (0) | Already binary variable |
| Parkinson's disease (self-reported) | Yes (1) or No (0) | Already binary variable |
| Dementia (self-reported) | Yes (1) or No (0) | Already binary variable |
| Arthritis (self-reported) | Yes (1) or No (0) | Already binary variable |
| Any other chronic disease (self-reported) | Yes (1) or No (0) | Already binary variable |
| Polypharmacy (self-reported) | >4 medications (1), ≤4 medications (0) | Recommended technique (Theou et al., 2013) |
| Body Mass Index | 18.5 to <25 (0), 25 to <30 (0.5), <18.5 or >equal to 30 (1) | Recommended technique (Chamberlain et al., 2016) |
| 6 m walk time (gait speed) | >10 seconds or physically unable (1), <10 seconds (0) | Recommended technique (Hoogendijk et al., 2017) |
| Able to stand up from a chair | Yes (1) or No (0) | Already binary variable |
| Grip strength (strongest hand and stratified by sex and BMI) | <5th percentile (1), 5th-20th percentile (0.5), >20th percentile (0) | Recommended technique (Theou et al., 2015) |
| Townsend Disability Scale | <5th percentile (1), 5th-20th percentile (0.5), >20th percentile (0) | Recommended technique (Theou et al., 2015) |
| Peak Expiratory Flow rate (stratified by sex) | <5th percentile (1), 5th-20th percentile (0.5), >20th percentile (0) | Recommended technique (Theou et al., 2015) |
| Forced expiratory volume (stratified by sex) | <5th percentile (1), 5th-20th percentile (0.5), >20th percentile (0) | Recommended technique (O Theou et al., 2015) |
| Depression (measured in the HADS) | 11 -21 (1), 8 – 10 (0.5), 0 – 7 (0) | Recommended technique (Zigmond & Snaith, 1983) |
| Anxiety (measured in the HADS) | 11 -21 (1), 8 – 10 (0.5), 0 – 7 (0) | Recommended technique (Zigmond & Snaith, 1983) |
| Mini-Mental State Examination | <10 (1), 11-17 (0.75), 18 – 20 (0.5), 20 – 24 (0.25), >24 (0) | Recommended technique (Searle et al., 2008) |
| Digit Symbol (measured in WAIS-III) | <5th percentile (1), 5th-20th percentile (0.5), >20th percentile (0) | Recommended technique (Theou et al., 2015) |
| Block Design (measured in WAIS-III) | <5th percentile (1), 5th-20th percentile (0.5), >20th percentile (0) | Recommended technique (Theou et al., 2015) |
| Verbal Fluency | <5th percentile (1), 5th-20th percentile (0.5), >20th percentile (0) | Recommended technique (Theou et al., 2015) |
| Matrix Reasoning (measured in WAIS-III) | <5th percentile (1), 5th-20th percentile (0.5), >20th percentile (0) | Recommended technique (Theou et al., 2015) |
| Reaction time test | <5th percentile (1), 5th-20th percentile (0.5), >20th percentile (0) | Recommended technique (Theou et al., 2015) |
| Delayed recall | <5th percentile (1), 5th-20th percentile (0.5), >20th percentile (0) | Recommended technique (Theou et al., 2015) |

Abbreviations: HADS - Hospital Anxiety and Depression scale; WAIS - Wechsler Adult Intelligence Scale

^a^Table is based on Table A1 in Welstead M, Muniz-Terrera G, Russ TC, et al. Inflammation as a risk factor for the development of frailty in the Lothian Birth Cohort 1936. *Exp Gerontol* 2020; 139: 111055.

**Supplementary Figure 2:** Johnson-Neyman plots of neighbourhood social deprivation slopes and their confidence bands depicting regions of significance conditional on age

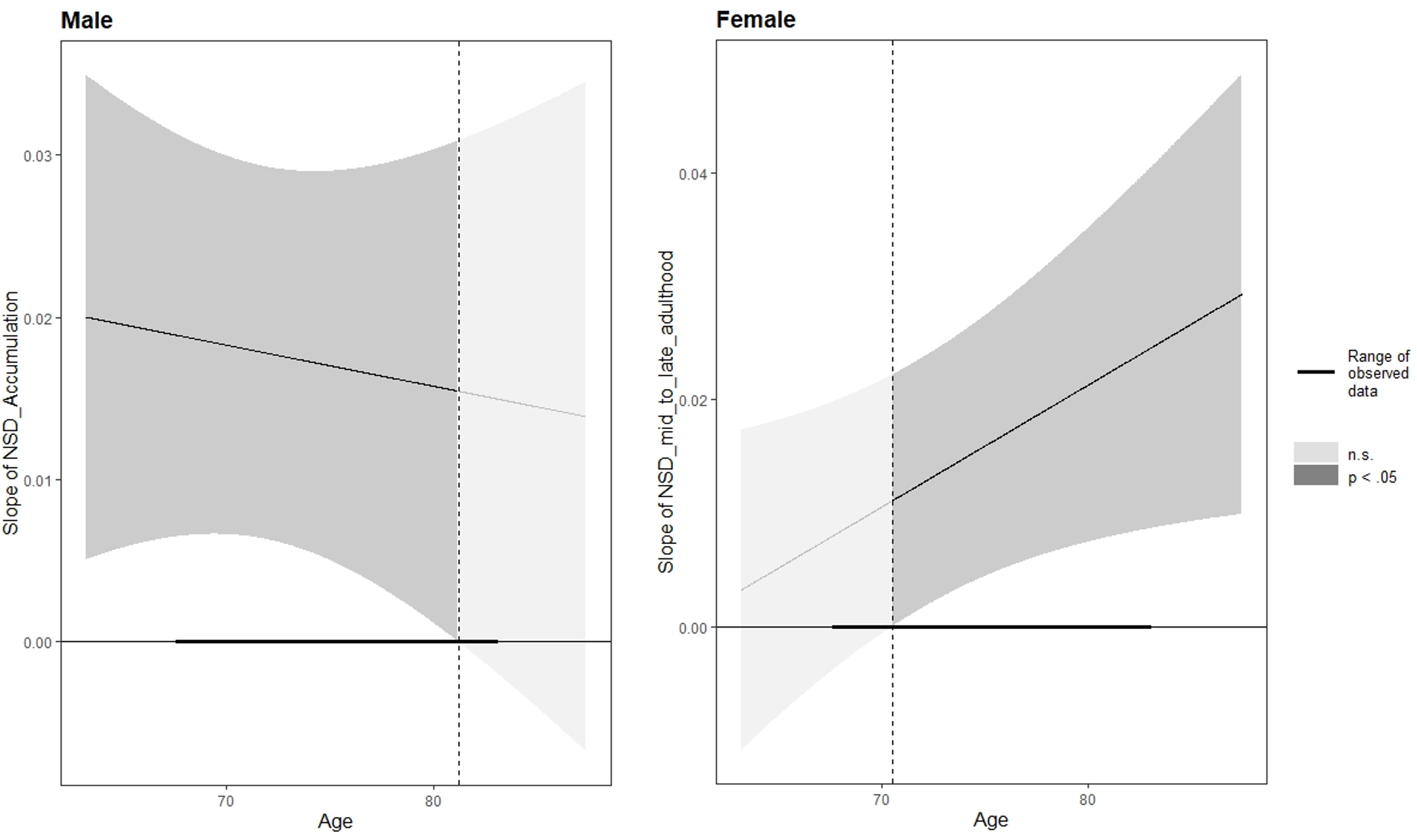

*Note***:** Darker grey areas indicate age intervals with significant, lighter grey areas with non-significant neighbourhood social deprivation and fraily associations; dark horizontal lines represent the range of observed data. Dotted verical lines depict the thresholds where significance changes in the male (81.23 years old) and female (70.48 years old) subsamples. Calculations are based on most appropriate life-course models for male (i.e. accumulation) and female (i.e. mid-to-late adulthood sensitive period) participants. Intervals were adjusted for false discovery rate.

**Supplementary Table 2**: Main models among Lothian Birth Cohort 1936 participants remaining in Edinburgh throughout their lives

|  | **Male (n=127)**  **Accumulation** | | | | **Female (n=120)**  **Mid-to-late adulthood sensitive period^a^** | | | |
| --- | --- | --- | --- | --- | --- | --- | --- | --- |
|  | b | 95% CI | *P*-value | β | b | 95% CI | *P*-value | β |
| **Frailty at age 70 (wave 1)** | | | | | | | | |
| Neighbourhood social deprivation | 0.015 | 0.001, 0.030 | 0.034 | 0.206 | 0.013 | -0.002, 0.027 | 0.081 | 0.189 |
| Age in years | 0.005 | -0.007, 0.018 | 0.406 | 0.072 | 0.015 | 0.003, 0.027 | 0.015 | 0.221 |
| Parental OSC (ref: I & II) | -0.012 | -0.044, 0.019 | 0.437 | -0.165 | -0.008 | -0.036, 0.021 | 0.601 | -0.114 |
| IQ at age 11 | -0.009 | -0.022, 0.004 | 0.182 | -0.116 | -0.020 | -0.032, -0.007 | 0.002 | -0.298 |
| Years spent in education | -0.022 | -0.039, -0.006 | 0.009 | -0.295 | 0.004 | -0.009, 0.017 | 0.542 | 0.059 |
| Childhood smoking (ref: no) | 0.004 | -0.023, 0.031 | 0.754 | 0.057 | 0.005 | -0.030, 0.040 | 0.772 | 0.077 |
| Adult OSC (ref: I & II) | 0.004 | -0.023, 0.032 | 0.761 | 0.057 | -0.015 | -0.039, 0.009 | 0.215 | -0.228 |
| Current smoking (ref: no) | 0.005 | -0.045, 0.055 | 0.841 | 0.068 | 0.006 | -0.036, 0.048 | 0.771 | 0.092 |
| **Frailty progression between age 70 and 82 (wave 1-5)** | | | | | | | | |
| *Fixed effects* |  | | | |  | | | |
| Neighbourhood social deprivation | 0.017 | 0.004, 0.030 | 0.012 | 0.193 | 0.018 | 0.005, 0.030 | 0.006 | 0.215 |
| Age in years | 0.029 | 0.024, 0.035 | 0.000 | 0.333 | 0.029 | 0.024, 0.034 | 0.000 | 0.353 |
| Parental OSC (ref: I & II) | -0.005 | -0.033, 0.023 | 0.739 | -0.055 | -0.004 | -0.029, 0.020 | 0.731 | -0.052 |
| IQ at age 11 | -0.008 | -0.019, 0.003 | 0.173 | -0.091 | -0.026 | -0.037, -0.015 | 0.000 | -0.316 |
| Years spent in education | -0.020 | -0.035, -0.005 | 0.009 | -0.232 | 0.003 | -0.007, 0.014 | 0.571 | 0.037 |
| Childhood smoking (ref: no) | 0.007 | -0.017, 0.031 | 0.576 | 0.079 | 0.010 | -0.019, 0.040 | 0.491 | 0.126 |
| Adult OSC (ref: I & II) | 0.001 | -0.024, 0.026 | 0.955 | 0.008 | -0.025 | -0.045, -0.004 | 0.019 | -0.302 |
| Current smoking (ref: no) | 0.013 | -0.032, 0.057 | 0.585 | 0.143 | 0.005 | -0.030, 0.041 | 0.772 | 0.064 |
| Neighbourhood social deprivation*Age | 0.001 | -0.004, 0.007 | 0.638 | 0.015 | 0.004 | -0.001, 0.009 | 0.129 | 0.050 |
| *Random effects* |  |  |  |  |  |  |  |  |
| Intercept, variance (SD) | 0.004 | (0.061) |  |  | 0.003 | (0.055) |  |  |
| Slope, variance (SD) | 0.001 | (0.024) |  |  | 0.000 | (0.022) |  |  |
| Residuals, variance (SD) | 0.001 | (0.038) |  |  | 0.001 | (0.038) |  |  |

*Note*: Regression coefficients (b) and 95% confidence intervals (CI) are displayed; we also provide fully standardized coefficients (β) to aid interpretation. Multivariate models are based on linear regression for frailty at age 70 and on linear mixed-effects regression with random intercepts and slopes for frailty progression; continuous predictors are mean centred and scaled. Abbreviations: OSC – occupational social class, SD – standard deviation

^a^ Models are adjusted for neighbourhood social deprivation in the previous developmental period (i.e. young adulthood).

**Supplementary Table 3**: Main models based on non-overlapping exposure and outcome measurements in the Lothian Birth Cohort 1936

|  | **Male (n=161)**  **Accumulation** | | | | **Female (n=161)**  **Mid-to-late adulthood sensitive period^a^** | | | |
| --- | --- | --- | --- | --- | --- | --- | --- | --- |
|  | b | 95% CI | *P*-value | β | b | 95% CI | *P*-value | β |
| **Frailty at age 70 (wave 1)** | | | | | | | | |
| Neighbourhood social deprivation | 0.017 | 0.004, 0.029 | 0.007 | 0.220 | 0.010 | -0.002, 0.022 | 0.100 | 0.150 |
| Age in years | 0.009 | -0.002, 0.019 | 0.113 | 0.115 | 0.012 | 0.002, 0.022 | 0.025 | 0.177 |
| Parental OSC (ref: I & II) | -0.019 | -0.044, 0.007 | 0.153 | -0.247 | -0.001 | -0.025, 0.023 | 0.933 | -0.015 |
| IQ at age 11 | -0.009 | -0.021, 0.002 | 0.108 | -0.126 | -0.018 | -0.029, -0.007 | 0.002 | -0.263 |
| Years spent in education | -0.020 | -0.034, -0.006 | 0.005 | -0.270 | 0.001 | -0.011, 0.013 | 0.816 | 0.021 |
| Childhood smoking (ref: no) | 0.011 | -0.013, 0.035 | 0.364 | 0.146 | 0.002 | -0.032, 0.036 | 0.900 | 0.032 |
| Adult OSC (ref: I & II) | 0.007 | -0.017, 0.031 | 0.580 | 0.089 | -0.018 | -0.041, 0.004 | 0.113 | -0.268 |
| Current smoking (ref: no) | -0.009 | -0.052, 0.034 | 0.688 | -0.117 | 0.004 | -0.032, 0.040 | 0.835 | 0.056 |
| **Frailty progression between age 70 and 82 (wave 1-5)** | | | | | | | | |
| *Fixed effects* |  | | | |  | | | |
| Neighbourhood social deprivation | 0.017 | 0.006, 0.029 | 0.004 | 0.197 | 0.016 | 0.005, 0.027 | 0.006 | 0.193 |
| Age in years | 0.029 | 0.024, 0.034 | 0.000 | 0.333 | 0.027 | 0.023, 0.032 | 0.000 | 0.328 |
| Parental OSC (ref: I & II) | -0.014 | -0.038, 0.010 | 0.257 | -0.159 | 0.007 | -0.015, 0.028 | 0.535 | 0.082 |
| IQ at age 11 | -0.009 | -0.020, 0.001 | 0.091 | -0.107 | -0.022 | -0.032, -0.012 | 0.000 | -0.265 |
| Years spent in education | -0.018 | -0.032, -0.005 | 0.008 | -0.209 | 0.001 | -0.009, 0.012 | 0.822 | 0.015 |
| Childhood smoking (ref: no) | 0.014 | -0.008, 0.036 | 0.214 | 0.162 | 0.005 | -0.025, 0.036 | 0.731 | 0.065 |
| Adult OSC (ref: I & II) | 0.004 | -0.019, 0.026 | 0.743 | 0.043 | -0.024 | -0.044, -0.004 | 0.022 | -0.285 |
| Current smoking (ref: no) | -0.002 | -0.043, 0.038 | 0.918 | -0.024 | 0.007 | -0.025, 0.040 | 0.659 | 0.088 |
| Neighbourhood social deprivation*Age | -0.001 | -0.006, 0.004 | 0.737 | -0.009 | 0.004 | -0.000, 0.009 | 0.053 | 0.053 |
| *Random effects* |  |  |  |  |  |  |  |  |
| Intercept, variance (SD) | 0.004 | (0.062) |  |  | 0.004 | (0.059) |  |  |
| Slope, variance (SD) | 0.001 | (0.023) |  |  | 0.000 | (0.021) |  |  |
| Residuals, variance (SD) | 0.001 | (0.037) |  |  | 0.001 | (0.039) |  |  |

*Note*: Regression coefficients (b) and 95% confidence intervals (CI) are displayed; we also provide fully standardized coefficients (β) to aid interpretation. Multivariate models are based on linear regression for frailty at age 70 and on linear mixed-effects regression with random intercepts and slopes for frailty progression; continuous predictors are mean centred and scaled. Abbreviations: OSC – occupational social class, SD – standard deviation

^a^ Models are adjusted for neighbourhood social deprivation in the previous developmental period (i.e. young adulthood).

**Supplementary Table 4**: Main models with categorical neighbourhood social deprivation variables in the Lothian Birth Cohort 1936

|  | **Male (n=161)**  **Accumulation** | | | | **Female (n=162)**  **Mid-to-late adulthood sensitive period^a^** | | | |
| --- | --- | --- | --- | --- | --- | --- | --- | --- |
|  | b | 95% CI | *P*-value | β | b | 95% CI | *P*-value | β |
| **Frailty at age 70 (wave 1)** | | | | | | | | |
| Neighbourhood social deprivation (ref: Low) |  |  |  |  |  |  |  |  |
| Medium | 0.014 | -0.014, 0.042 | 0.318 | 0.186 | 0.018 | -0.008, 0.044 | 0.172 | 0.268 |
| High | 0.040 | 0.011, 0.069 | 0.007 | 0.532 | 0.007 | -0.021, 0.036 | 0.607 | 0.110 |
| Age in years | 0.009 | -0.002, 0.020 | 0.108 | 0.117 | 0.012 | 0.002, 0.023 | 0.025 | 0.178 |
| Parental OSC (ref: I & II) | -0.019 | -0.044, 0.007 | 0.156 | -0.248 | 0.002 | -0.022, 0.026 | 0.870 | 0.029 |
| IQ at age 11 | -0.009 | -0.021, 0.003 | 0.132 | -0.119 | -0.019 | -0.030, -0.008 | 0.001 | -0.276 |
| Years spent in education | -0.022 | -0.036, -0.008 | 0.002 | -0.291 | -0.000 | -0.012, 0.011 | 0.941 | -0.006 |
| Childhood smoking (ref: no) | 0.009 | -0.015, 0.033 | 0.448 | 0.122 | 0.005 | -0.029, 0.040 | 0.758 | 0.080 |
| Adult OSC (ref: I & II) | 0.005 | -0.020, 0.029 | 0.709 | 0.061 | -0.016 | -0.039, 0.007 | 0.162 | -0.237 |
| Current smoking (ref: no) | -0.009 | -0.052, 0.034 | 0.677 | -0.122 | 0.001 | -0.035, 0.038 | 0.953 | 0.016 |
| **Frailty progression between age 70 and 82 (wave 1-5)** | | | | | | | | |
| *Fixed effects* |  | | | |  | | | |
| Neighbourhood social deprivation (ref: Low) |  |  |  |  |  |  |  |  |
| Medium | 0.021 | -0.006, 0.048 | 0.124 | 0.245 | 0.031 | 0.006, 0.055 | 0.016 | 0.369 |
| High | 0.035 | 0.007, 0.062 | 0.017 | 0.395 | 0.032 | 0.005, 0.058 | 0.019 | 0.384 |
| Age in years | 0.029 | 0.021, 0.037 | 0.000 | 0.332 | 0.019 | 0.011, 0.027 | 0.000 | 0.232 |
| Parental OSC (ref: I & II) | -0.014 | -0.039, 0.010 | 0.251 | -0.163 | 0.010 | -0.012, 0.031 | 0.376 | 0.117 |
| IQ at age 11 | -0.008 | -0.019, 0.002 | 0.130 | -0.097 | -0.023 | -0.033, -0.013 | 0.000 | -0.275 |
| Years spent in education | -0.021 | -0.034, -0.008 | 0.002 | -0.237 | -0.001 | -0.011, 0.010 | 0.918 | -0.007 |
| Childhood smoking (ref: no) | 0.013 | -0.010, 0.035 | 0.260 | 0.149 | 0.008 | -0.023, 0.039 | 0.609 | 0.097 |
| Adult OSC (ref: I & II) | 0.003 | -0.020, 0.026 | 0.798 | 0.034 | -0.022 | -0.042, -0.002 | 0.032 | -0.266 |
| Current smoking (ref: no) | -0.001 | -0.042, 0.039 | 0.946 | -0.016 | 0.004 | -0.029, 0.037 | 0.804 | 0.050 |
| Neighbourhood social deprivation*Age (ref: Low*Age) |  |  |  |  |  |  |  |  |
| Medium*Age | 0.005 | -0.007, 0.017 | 0.400 | 0.057 | 0.007 | -0.004, 0.017 | 0.233 | 0.080 |
| High*Age | -0.004 | -0.015, 0.007 | 0.483 | -0.046 | 0.016 | 0.005, 0.027 | 0.005 | 0.190 |
| *Random effects* |  |  |  |  |  |  |  |  |
| Intercept, variance (SD) | 0.004 | (0.063) |  |  | 0.004 | (0.059) |  |  |
| Slope, variance (SD) | 0.001 | (0.023) |  |  | 0.000 | (0.020) |  |  |
| Residuals, variance (SD) | 0.001 | (0.037) |  |  | 0.001 | (0.039) |  |  |

*Note*: Regression coefficients (b) and 95% confidence intervals (CI) are displayed; we also provide fully standardized coefficients (β) to aid interpretation. Multivariate models are based on linear regression for frailty at age 70 and on linear mixed-effects regression with random intercepts and slopes for frailty progression; continuous predictors are mean centred and scaled. Abbreviations: OSC – occupational social class, SD – standard deviation

^a^ Models are adjusted for neighbourhood social deprivation in the previous developmental period (i.e. young adulthood).
